# Association Between Acromion Morphology and Non-Traumatic Rotator Cuff Injury Among Young Adult Filipinos in a Tertiary Hospital: Cross-sectional Study

**DOI:** 10.1101/2025.07.24.25332122

**Authors:** Terence Aaron L. Burgo, Brian Andrich L. Pollo

## Abstract

**Introduction:** Rotator cuff injury (RCI) is among the most common shoulder pathologies, significantly affecting quality of life. Acromial morphology has been implicated as a predisposing factor, but limited local data exist. The Bigliani-Kitay classification defines four acromion types: flat (I), curved (II), hooked (III), and convex (IV). MRI is the preferred modality for assessing both acromial morphology and RCI.

**Objective:** To assess the association between acromion morphology and RCI among patients undergoing shoulder MRI at Makati Medical Center, a tertiary hospital in the Philippines.

**Methods:** A retrospective review of shoulder MRI scans performed from January to December 2019 was conducted. Patients with shoulder pain were included as cases. Acromion morphology and presence of RCI (tendinosis, partial, or full-thickness tears) were documented. Images were interpreted by radiologists accredited by the Philippine College of Radiology.

**Results:** Of 132 patients (mean age: 31.2 ± 5.4 years), 65.9% were male. The period prevalence of RCI was 58.3% (n = 77). There was no significant sex-based difference in RCI prevalence (*p* = 0.4036). RCI was significantly associated with Type II (70.5%, *p* < 0.00001) and Type III (100%, *p* = 0.00158) acromion morphologies. No significant association was found for Types I and IV.

**Conclusion:** Type II and Type III acromial morphologies are significantly associated with higher prevalence of RCI. Recognition of these morphologies on MRI may help identify patients at increased risk and allow for early intervention.

**Advances in knowledge:**

- Confirms a statistically significant association between Type III (hook-shaped) acromion and rotator cuff injury (RCI) in a Filipino young adult population.
- While associated with RCI as well, type II acromion is most prevalent in the study population.
- This is among the first studies in the Philippines to characterize acromial types in relation to documented RCI.

**Application to patient care:**

- Early identification of type III acromion may support clinical suspicion for rotator cuff pathology, improving diagnostic accuracy.
- Patients with type III acromion may benefit from earlier referral to physical therapy or orthopedic evaluation.
- Incorporating acromial morphology in radiology reports may enhance multidisciplinary communication between radiologists, primary care providers, and orthopedic surgeons.

## Introduction

Rotator cuff injuries (RCI) are among the most common shoulder pathologies, yet their precise etiology remains incompletely understood.^1^ While both intrinsic factors (*e.g.,* age-related degeneration, reduced vascularity, and collagen abnormalities) and extrinsic factors (*e.g.,* subacromial impingement and repetitive overhead activity) have been implicated, relatively few studies have focused on the role of specific acromion morphologies in predisposing individuals to RCI. Of these, the hook-shaped type III acromion has been most consistently associated with rotator cuff pathology.^2^

Mechanical impingement is a leading extrinsic cause of rotator cuff injury, implicated in up to 95% of tears.^3^ It results from repetitive compression of the tendons under the acromion, contributing to tendon degeneration. Acromioplasty, which addresses this impingement, has become increasingly common. Early diagnosis and management of mechanical factors are associated with better outcomes, emphasizing the need to identify structural contributors, such as acromial morphology.

Acromial morphology is a key anatomical determinant of subacromial impingement.^4,5^ The most widely used system for classifying acromion types is the Bigliani classification, later refined by Kitay et al., which describes four morphological types based on the shape of the inferior acromial surface: flat (Type I), curved (Type II), hooked (Type III), and convex (Type IV).^6^ Magnetic Resonance Imaging (MRI) has since emerged as the most accurate modality for assessing both acromion shape and rotator cuff integrity.

Several studies have examined the relationship between acromion type and impingement or RCI. While many report an association between Type III (hooked) acromion and non-traumatic RCI,^2,7–11^ others have found no significant relationship.^12,13^ Such conflicting results demonstrate the ongoing uncertainty in the role of acromial morphology in RCI pathogenesis. Notably, nearly all of these studies were conducted among middle-aged and older adults, leaving a gap in understanding of how acromial morphology relates to RCI in younger populations. Moreover, there is a lack of local data from the Philippines on this issue. Hence, the primary objective of this study was to assess the association between acromial morphology and non-traumatic rotator cuff injury among young adult Filipinos (<40 years old) using MRI-based classification.

RCIs are known to reduce quality of life to levels comparable with chronic conditions such as heart failure, diabetes, and depression. Treatment costs can accumulate rapidly, and delays in appropriate management can result in poorer outcomes. Early identification of anatomical risk factors, particularly in younger adults, may aid in preventive strategies and more tailored treatment approaches.^11^

## Methods

This cross-sectional study retrospectively reviewed the MRI results of all patients (ages 25 to 50 years) of Makati Medical Center who underwent shoulder MRI from January 1, 2019 to December 31, 2019 as interpreted by radiologists accredited by the Philippine College of Radiology and fellows of the CT-MRI society of the Philippines. This study included as cases all patients with RCI, either tendinosis, partial or full thickness. We excluded individuals with (1) previous shoulder surgery, (2) fractures and/or dislocation, (3) infections or tumors of the shoulder, and (4) presence of acromial spurs. Asymptomatic patients with intact rotator cuff were also included to serve as control.

All scans were performed using a 1.5-Tesla MRI scanner (Siemens). A shoulder array coil was used. The patients were positioned supine with their arms on the sides of their body in partial external rotation. Coronal oblique, sagittal oblique and axial images were obtained. The coronal oblique plane were selected parallel to the course of the supraspinatus tendon for ideal image acquisition of the tendon. The sagittal oblique image was used to identify acromion morphology.

The main outcome was the presence of rotator cuff injury was determined by radiologic report. Complete rotator cuff tears were identified as a hyperintense signal area within the tendon on T2W and fat-suppressed images, seen in two planes. Complete tears are described as T2/PD signals extending from the articular or bursal surface, which most commonly affect the supraspinatus tendon. Fluid signal within the tendon defect is highly suggestive of rotator cuff injury.^14^

Partial tears were defined as T2/PD signals that extend in either, not both, the bursal or articular surface. Partial tears are also seen within the tendon substance, hence the term, intrasubstance partial tear. Radiologists use tendinosis as an all-encompassing term to indicate all tendon pathology. Also, they may use it to suggest a chronic tendon disease that does not resolve.^15^

The primary exposure was acromion morphology, classified using the Bigliani system by a single radiologist (T.B.). Information on age and sex were collected as well. MRI reports and images of all patients who underwent shoulder MRI were retrieved from the Novarad radiology information system (NovaRIS) and picture archiving and communication system (NovaPACS) of the Department of Radiology. The data were tabulated using Microsoft Excel-based data abstraction tool.

The sample size was computed with 95% confidence level, an acceptable margin of error at 5%, an estimated prevalence of rotator cuff injury in certain acromion morphology of 56%.^12^ The computation for sample size suggested a need for at least 132 patients.

MRI reports and images of all patients who underwent shoulder MRI were retrieved from the NOVARAD RIS and PACS system of the Department of Radiology. The data were tabulated using Microsoft Excel-based data abstraction tool.

Descriptive statistics were used to describe the demographic characteristics (age and sex) of the participants. Qualitative and quantitative data were numerically expressed as frequencies, proportions and means ± standard deviations (SD).

The prevalence of rotator cuff injury according to sex and acromion morphology were determined. For each acromion type, a 2×2 contingency table was constructed comparing the number of patients with the given type versus all others, stratified by RCI status. Fisher’s Exact Test was used for all comparisons due to the small sample sizes, particularly for Types III and IV. A *p*-value less than 0.05 was considered statistically significant. All statistical tests were conducted in R (v.4.5.0).

## Results

A total of 155 Shoulder MRI studies of young adult patients were reviewed and 132 of them were included in the study due to presence of one of the following: injury of the rotator cuff, either tendinosis, partial or complete tear with no history of previous shoulder surgery, no associated fractures nor dislocation, infections or tumors of the shoulder nor presence of degenerative changes. Of the 155 studies, 20 were not included due to a history of trauma with fracture and/or dislocation; 5 were also not included due to the presence of degenerative changes as stated in the exclusion criteria above.

The mean (± SD) age of the participants is 31.22 ± 5.42. Of the 132 participants included in the study, there was a higher proportion of males compared to females (Table 1). The overall period prevalence of rotator cuff injury was 58.33% (n = 77). There was higher prevalence of RCI among males, as opposed to females. There was no significant difference between the prevalence of rotator cuff injury among male subjects when compared to that of female subjects (p = 0.4036).

**Table 1.** Association between sex and presence of rotator cuff injury.

|  | (+) Rotator Cuff Injury<br>(n = 77) | (-) Rotator Cuff Injury<br>(n = 55) | Total<br>(n = 132) |
| --- | --- | --- | --- |
| Male | 60.92% (n = 53) | 39.08% (n = 34) | 100% (n = 87) |
| Female | 53.33% (n = 24) | 46.67% (n = 21) | 100% (n = 45) |

To determine which acromial shape was most associated with injury, we compared RCI incidence across types. Among all acromion types, Type II acromion, the most common variant, showed the strongest association with RCI (*p* < 0.00001). Type III was also linked to higher RCI rates (*p* = 00158). Although Type III acromion was present in only 5 patients, it was associated with a 100% rate of rotator cuff injury. Type II acromion, the most common variant, was also linked to higher RCI rates (*p* < 0.00001), possibly due to its curved morphology. Fisher’s exact test did not demonstrate a significant relationship between rotator cuff injury and the presence of type I acromion (*p* = 0.2041) or type IV acromion (*p* = 0.5287). Detailed frequencies are shown in Table 2.

**Table 2.**
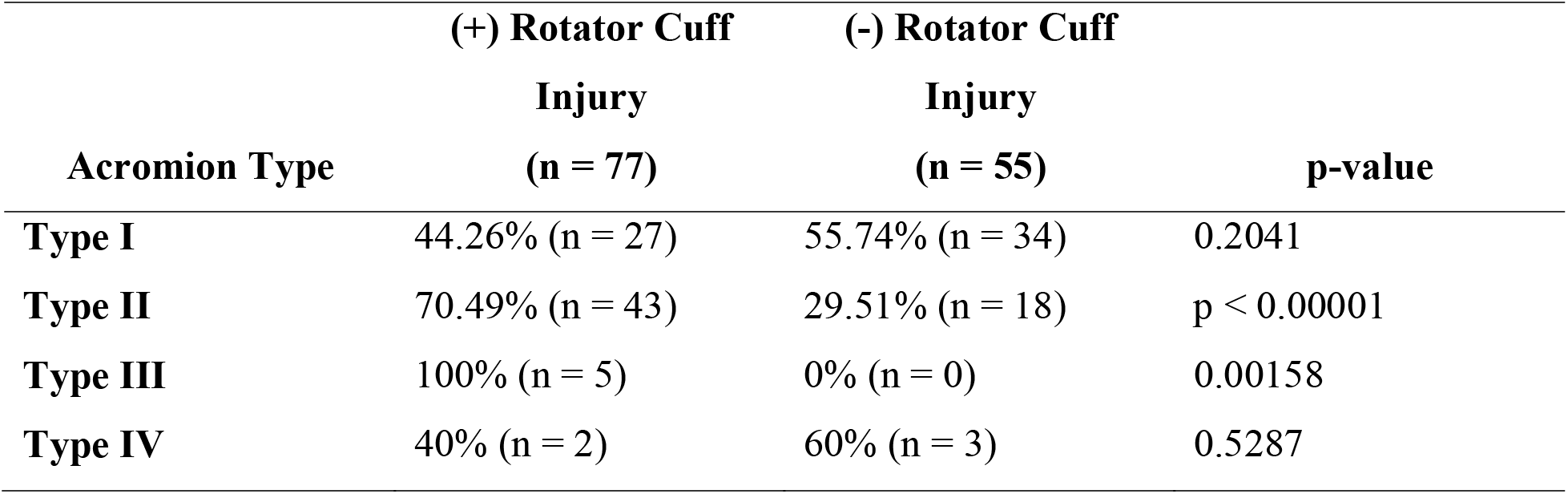
Association between acromion morphology and presence of rotator cuff injury.

## Discussion

The most significant finding in this study is the significant association between acromial morphology and non-traumatic RCI. Only ∼3% of the subjects had Type III acromion and although rare, all of them were associated with RCI (*p* = 0.00158) compared to other acromion types. This results align with systematic review findings that Type□III morphology nearly triples the odds of rotator cuff tear compared to Types I–II (OR =□2.82, *p* =□3e^-6^).^11^ This association is probably due to shape of Type III acromion described as being hooked. A hooked acromion differs from the Type I and II due to its increased anterior and inferior extension of the acromial bony process, which is thought to decrease the subacromial space, thereby increasing the likelihood of extrinsic degeneration of the rotator cuff tendons.^11^

Type II acromion was also found to be associated with increased occurrence of RCI (*p* < 0.00001) which may be due to its curved shape which can also narrow the subacromial space to some extent. However, being the most common type, Type II acromion can therefore significantly instigate a high incidence rate of RCI among all of the acromion types and can notably influence the causal relationship between this certain type and incidence of injury.^16^

Majority of the subjects were male but there is no significant difference in the incidence of RCI between sexes. Studies in different populations similarly report no consistent sex-based differences in acromial morphology among rotator cuff tear cases.^17,18^ Also, this study also shows similar results to studies conducted with different age cohorts, suggesting that Type□III acromion is a risk marker regardless of age. For instance, in a multivariable analysis of surgical cases, acromial morphology (especially type III) remained significantly associated with RCI even after stratification by age.^19^ But it is unknown whether these relationships are causal, or simply predictive in nature. Cross-sectional design limits causal inferences. Some investigators have found no significant correlation between acromion type and cuff tear, reinforcing uncertainty in the impingement hypothesis.^12,17,20^

These findings support using MRI-based acromial classification to identify individuals at elevated risk for non-traumatic RCI. Early recognition through imaging may guide targeted intervention, such as referral to physical therapists, physiatrists or orthopedic surgeons.

This study had some limitations. The analysis was not adjusted for many other extrinsic and intrinsic factors affecting the rotator cuff tendons’ risk of injury. Also, the relationship of the severity/grade of rotator cuff injury and the acromion types was not taken into consideration. Future prospective studies incorporating multivariate modeling and grading of tendon pathology may be needed to clarify causality.

## Conclusion

This study corroborates previous findings that type III acromion, although rare, is associated with a high incidence of non-traumatic rotator cuff injury. Type II acromion was also associated with RCI but it also has the highest prevalence. No significant association was found between sex and rotator cuff injury. Identifying those patients at risk may potentially benefit from corrective or preventive measures to help prevent future injury. Future studies may benefit by examining a greater number of factors and including a better distribution of acromion types.

## Data Availability

All data produced are available online at

https://doi.org/10.7910/DVN/IEVMZ1

## ETHICS COMMITTEE APPROVAL

Ethical approval was waived by the IRB of Makati Medical Center

## AUTHORS CONTRIBUTIONS STATEMENT

Terence Burgo was responsible for Investigation, Resources, Supervision, Writing – Original Draft, and Writing – Review & Editing. Brian Pollo contributed to contributed to Conceptualization, Data Curation, Literature Review, Writing – Original Draft, and Writing – Review & Editing. Both authors approved the final version of the manuscript.

